# Transcatheter closure of ventricular septal defects by exclusive transvenous antegrade cannulation from the right ventricle

**DOI:** 10.1101/2024.05.28.24308078

**Authors:** Pramod Sagar, Puthiyedath Thejaswi, Ishita Garg, Kothandam Sivakumar

## Abstract

**Background:** Transcatheter closure(TCC) of perimembranous ventricular septal defects(PM-VSD) and intraconal defects routinely involves either arteriovenous loop formation or direct retrograde device deployment from its left ventricular(LV) end. An arterial access is always taken for a left ventriculogram. Direct antegrade cannulation of the defect from the right ventricle for TCC avoids complications associated with femoral arterial access and arteriovenous loop formation.

**Methods:** Feasibility of elective antegrade cannulation for TCC of PM-VSD, intraconal VSD and postoperative residual VSD was retrospectively studied over five years from 2019-2023. Echocardiographic VSD measurements guided the device selection rather than left ventriculographic measurements. Predictors for successful antegrade cannulation and transvenous device deployment were analyzed.

**Results:** Antegrade cannulation was electively attempted in 116/163(71%) TCC VSD closure procedures. The proportion of cases where this antegrade cannulation was electively employed progressively increased from 26% of interventions performed in 2019 to 93% in 2023. The median age of the study cohort that included 24 infants was 55 months (range 1-636 months) and the indexed VSD size was 9.2 mm/m^2^ (range 1.7-43.3 mm/m^2^). Two-thirds of patients had varying degrees of pulmonary arterial hypertension. Antegrade cannulation was successful in 97(83.6%) patients. In the remaining 19 patients, retrograde cannulation from LV aided TCC. There were no deaths or need for pacemaker implantation. Overall procedural success of TCC in this cohort was 99.1%.Device embolization with tricuspid chordal tear led to procedural failure in one patient. Four other device embolizations were managed successfully by transcatheter retrieval and closure with an upsized device. Young age, small body size, large VSD size were significantly associated with successful antegrade cannulation.

**Conclusions:** Antegrade cannulation and TCC was feasible in majority of the procedures, especially in small patients and large defects. This strategy simplified the procedure without arterial access and might replace the routine retrograde device delivery and AV loop formation.

Graphical abstract
An exclusive transvenous antegrade defect cannulation from the right ventricle would avoid complications due to arterial access and arteriovenous railroad formation, simplify procedure and allow assessment of aortic valve before device release.

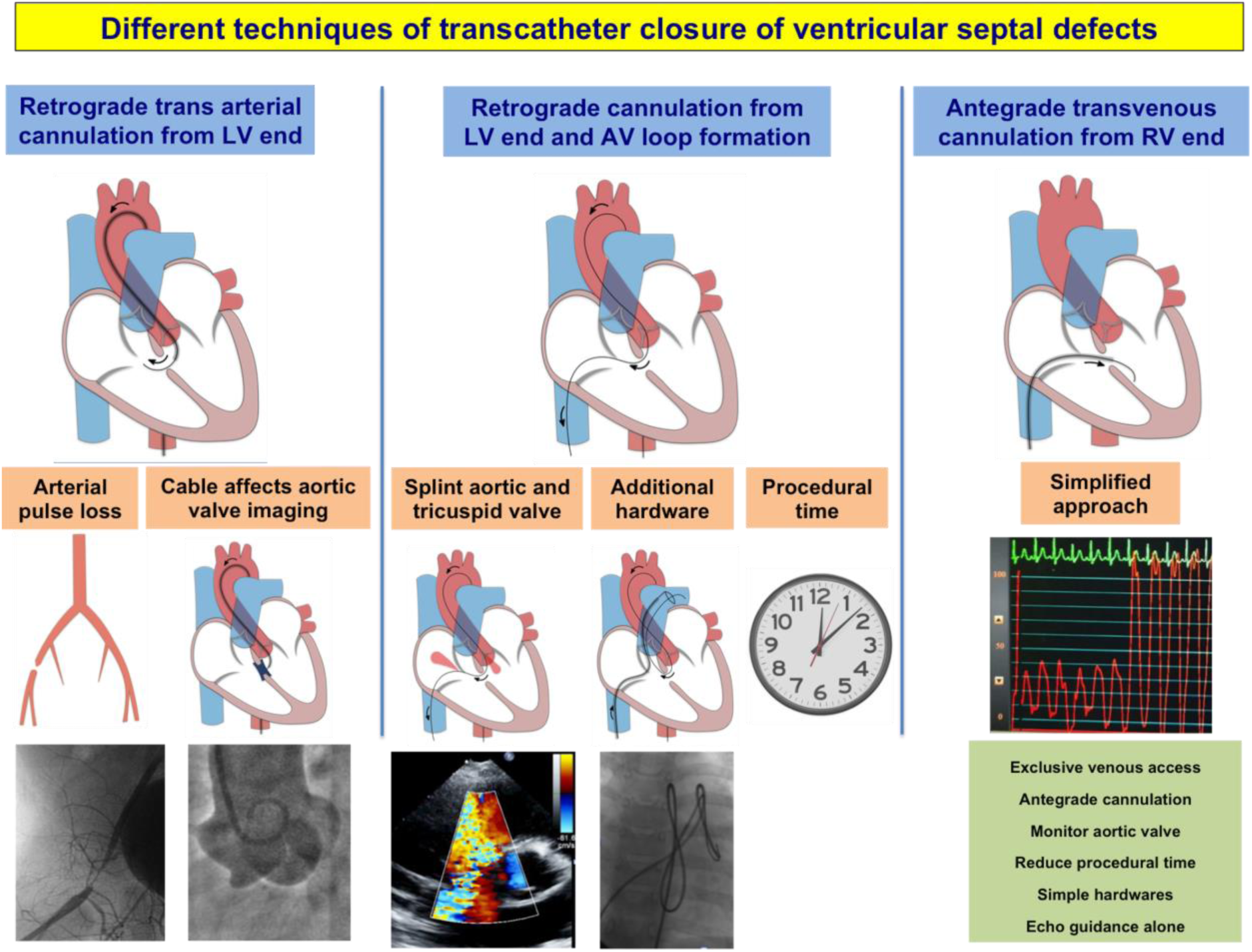

**What is already known:**

- Transcatheter closure of ventricular septal defects around the membranous septum is increasingly performed using soft occluder devices with results comparable to surgery.
- Conventional approach involves a retrograde trans-arterial cannulation of the defect or arteriovenous loop formation.

**What this study adds:**

- Transvenous antegrade cannulation and device deployment is an alternative attractive technique.
- As this avoids arterial access, additional hardware needed for arteriovenous loop formation and allows intraprocedural monitoring of aortic valve, this might replace the conventional techniques in future.

## Introduction

Transcatheter closure(TCC) of perimembranous ventricular septal defects(PM-VSD) re-emerged as an alternative to surgery in the last decade. Availability of soft, low-profile off-label devices with low radial and clamp force instead of the previous asymmetric occluders led to this resurgence.^[1–4]^ TCC achieved comparable results to surgery despite close proximity of the VSD to the anteroseptal commissure of the tricuspid valve, non coronary cusp(NCC) - right coronary cusp(RCC) commissure of the aortic valve and conduction tissue.^[5–9]^ These encouraging results led to adoption of device closure even in infants.^[10]^ Intraconal VSDs situated below the RCC of the aortic valve, common in oriental population were also recently considered amenable for TCC using such soft devices.^[11,12]^ The choice among the various off-label device designs were dependent on the anatomical variations between the different types of these defects.^[2,13]^

TCC was routinely performed after an initial left ventricular(LV) angiography from an arterial access followed by passage of a guide wire to cross the defect from the LV to the right ventricle(RV). The occluder was retrogradely deployed after exchanging a coronary guide catheter or a delivery system from this arterial access.^[12,14]^ Otherwise the guidewire was snared from the femoral venous end to establish an arteriovenous(AV) loop. A similar delivery catheter advanced into the aorta or the LV over this AV loop would allow a transvenous deployment of an occluder. Another alternative approach involved direct cannulation of the VSD from the RV end to advance a delivery catheter and deploy an occluder from the venous end. A similar exclusive use of venous access without an arterial puncture made a paradigm shift in TCC of patent arterial ducts.^[15–17]^ PM-VSD and intraconal defects were amenable for this antegrade cannulation due to their anatomical location. Significant advantages of this strategy were (i) unhindered intraprocedural monitoring for aortic regurgitation without any rigid cables across the valve, (ii) avoiding arterial vascular complications by using a single venous access, (iii) simplified procedure without AV loop formation that needed additional hardware, (iv) preventing hemodynamic compromise in young infants by splinting aortic and tricuspid valve leaflets through the AV loop and (v) finally reducing the procedural duration.

Use of venous access for antegrade cannulation of PM-VSDs tested in a swine model under magnetic resonance fusion imaging was simpler, faster and reduced radiation exposure compared to a conventional retrograde approach.^[18]^ Antegrade cannulation of these VSDs were occasionally performed by various operators during TCC of these VSDs but the numbers were limited. Moreover, the method of crossing the defect was not uniform and no studies evaluated the feasibility of antegrade cannulation, success rate of crossing, efficacy and safety.^[19–21]^ Analysis of feasibility and safety of antegrade crossing of PM-VSDs, intraconal and post-surgical residual defects would determine the factors influencing successful antegrade cannulation.

### Methodology

This prospective, single-centre study evaluated the feasibility of antegrade cannulation during transcatheter VSD closures performed between January 2019 and December 2023. Informed written consent was obtained for the intervention. Institutional ethics committee approved the study and anonymized reporting of the results.

#### Pre procedural assessment

All cases of PM-VSDs, intraconal VSDs and post-surgical residual VSDs were evaluated for indication for closure. Symptomatic patients and those with transthoracic echocardiographic evidence of left-heart volume overload were considered for TCC.

Subcostal, apical and parasternal views classified the type of VSD(A-D) based on the extent of aortic margin, presence of membranous septal aneurysm or aneurysm formed by septal tricuspid leaflet chordal attachments.^[2,13]^ The size of the defect was measured from the narrowest exit on the RV side of the defect. Whenever this orifice was oval on orthogonal views, the larger dimension was considered as the size of the defect. Device closure were not considered if RCC or NCC prolapse was significant with more than grade 2 aortic regurgitation(AR) and if the associated cardiac defects were not amenable for transcatheter correction.

#### Evolution of the procedure

All patients for whom an initial attempt was made to antegradely cannulate the defect and achieve a transvenous device closure were included in this study. Patients planned for an elective retrograde transarterial cannulation or elective AV loop formation were excluded. In the initial part of the study, PM-VSD without aortic margin needing intraprocedural echocardiographic monitoring of the aortic valve for regurgitation and small patients at risk of arterial access complications were considered for transvenous antegrade crossing due to the previously mentioned advantages. As experience of antegrade cannulation improved, few intraconal VSDs and post-surgical residual VSDs in the perimembranous location were also electively planned for antegrade crossing from the RV in the later part of the study. In the last two years, each and every case of PM-VSD, intraconal VSD and even post device closure residual defects were included for antegrade cannulation strategy.

#### Anaesthesia and access

Children less than 12 years underwent the procedure under intravenous conscious sedation with ketamine, fentanyl or midazolam, while adolescents and adults received local anaesthesia only. Femoral venous access was used in all cases. Arterial access was used in few earlier patients for performing LV angiogram. In recent times, arterial access was avoided and device closure was performed only on echocardiographic guidance. Arterial access was taken only when a very large VSD with severe pulmonary hypertension warranted a hemodynamic assessment of shunt ratio and pulmonary vascular resistance, or when antegrade cannulation failed. Unfractionated heparin was administered in a dose of 100 U/Kg after obtaining the venous access.

#### Technique of antegrade crossing

A standardized technique was adopted for crossing the VSD antegrade from the RV. A transvenous 4F or 5F Judkins right coronary catheter was advanced into the pulmonary artery(PA). While crossing the tricuspid valve, a loop of the guidewire or catheter was advanced through the central valve orifice ensuring that the tip of the catheter did not pass through the interchordal spaces. After connecting the catheter hub to fluid-filled pressure transducer, it was slowly withdrawn from the PA towards the right ventricular inflow. The catheter tip was pointed posteriorly towards the interventricular septum on a shallow left anterior oblique(45°) and cranial(30°) fluoroscopic projection in PM-VSD during this slow withdrawal. A lateral projection with cranial(30°) angulation was used in intraconal defects. A gentle clockwise and counter-clockwise torque of the catheter would give a cranial or caudal angulation respectively to point the catheter tip towards the VSD. A change in pressure waveform from RV pattern to LV pattern would indicate the proximity of the catheter tip to the VSD.(Figure 1) At this point, either the catheter was gently advanced to cannulate the VSD or a 0.035” angled hydrophilic Glidewire(Terumo corporation, Tokyo, Japan) was used to cannulate the VSD to enter the LV. Among the different makes of right Judkins catheters, Optitorque(Terumo corporation, Tokyo, Japan) with a less acute primary curve was preferred in infants compared to the sharp curve of Infiniti(Cordis, Miami Lakes, FL) preferred in older patients. As a more acute curve was desired in adults, internal mammary catheters were used in this group.

**Figure 1:**
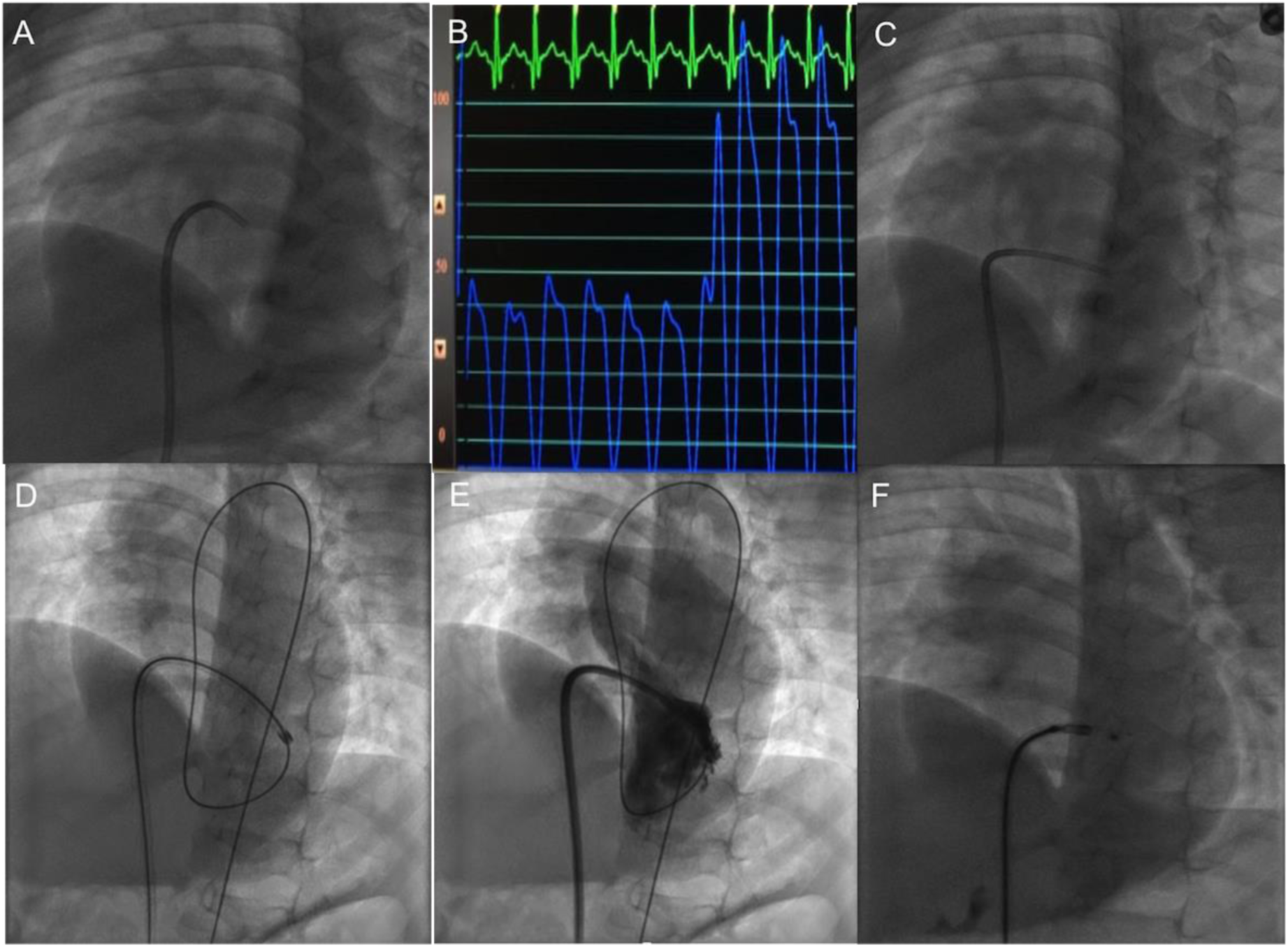
Antegrade cannulation. Tip of a Judkins right coronary catheter facing posteriorly towards the ventricular septum in left anterior oblique cranial projection(A) detects a change in pressure from right ventricular to left ventricular form(B) when it abuts the ventricular septal defect. Advancing the catheter to cannulate the defect(C) followed by an exchange to a Mullins sheath over the guidewire(D) allows a left ventricular angiogram from the sidearm of the sheath(E) before a transvenous direct device deployment(F).

#### Advancing the delivery system

After cannulating the VSD and entry into the LV, the catheter or Glidewire was exchanged to a Launcher right coronary guide catheter(Medtronic, Santa Rosa, CA) or a Mullins sheath(Cook Medical, Bloomington, IN). When the support from the Glidewire was considered inadequate for advancing a Mullins sheath, it was exchanged for a 260cm long 0.035” Amplatzer Super Stiff wire(Boston scientific, Natick, MA) with 6cm floppy J-tip. Once the delivery catheter or sheath was advanced into a stable position in the LV, contrast was injected with hand to obtain a left ventriculogram to identify the location of the VSD and the aortic valve to guide the device delivery. No attempts were made to measure the defect size in this angiogram, as all decisions regarding the defect measurements were exclusively based on echocardiography.

#### Choice of device design and size

The device design was chosen based on the type of PM-VSD as described earlier.^[2,13]^ The most commonly used devices included Amplatzer duct occluder(ADO), Amplatzer duct occluder-II(ADO-II) or Amplatzer muscular VSD occluder(Abbott vascular, Golden Valley, MN) and HeartR duct occluder or Konar-MF VSD occluder(Lifetech Scientific, Shenzhen, PRC). ADO-II or Konar-MF were soft and preferred for closing intraconal VSD. The devices were chosen 0 to 2 mm larger than the defect size measured by transthoracic echocardiography. Other devices such as Piccolo(Abbott vascular, Golden Valley, MN) and Memopart muscular VSD occluder(Lepu Medical, Beijing, PRC) were rarely used. Oversizing was avoided in very small infants and defects without aortic margin.

#### Device deployment

The devices were deployed from the transvenous delivery catheter. Pressure from the tip of the delivery catheter was monitored through the side-arm of the Mullins sheath or using a Y-connector for the guide catheters. This allowed precise LV and RV disc deployment when the pressure waveform transitioned from LV pattern to RV pattern during a progressive withdrawal of the delivery sheath. Additional guidance was also obtained from a transthoracic echocardiogram. Echocardiographic guidance was crucial when the LV and RV pressures were similar in patients with severe hyperkinetic pulmonary hypertension. Transesophageal echocardiogram was not used. The appropriateness of the device position, its stability and presence of residual flows, occurrence of AR, if any, were assessed by transthoracic echocardiography before releasing the device. If new onset AR was noted, the device was either repositioned or changed to get acceptable results.

#### Failure of antegrade cannulation

Whenever antegrade crossing failed, an arterial access was taken and the VSD was crossed retrogradely in the conventional manner. Subsequently the device was deployed either retrograde from the same arterial catheter or antegradely from a transvenous sheath placed after formation of an AV loop.

#### Post-procedural care and follow-up

Haemostasis was achieved by manual compression. Patients were monitored for rhythm disturbances, valvular or vascular complications for 1-2 days after the procedure. No anticoagulation or antiplatelets were advised on discharge. Clinical, electrocardiographic and echocardiographic evaluation were performed at 1-month, 6-month follow-up and yearly thereafter. At each follow up visit, rhythm abnormalities, presence and severity of valvular regurgitation, residual leaks and their significance were assessed.

#### Statistical analysis

Statistical analysis was performed using IBM SPSS Statistics version-25(IBM Corp, Armonk, NY). Categorical variables were expressed as a frequency or a percentage. Continuous variables were presented as median with range or mean with standard deviation as appropriate. Chi-square test was used for categorical variables and Mann-Whitney U test for continuous variables during assessment of predictors of failed antegrade cannulation. Wilcoxon signed rank test was used to evaluate the effect of VSD closure on Weight z-score, LV dimensions and severity of valvular regurgitations on follow up. A value for p-value less than 0.05 was considered statistically significant.

## Results

Among a total of 163 cases of PM-VSDs, intraconal and post-surgical residual VSD in membranous location that underwent device closure during the 5-year study period, an elective attempt was made to antegradely cannulate the VSD from the RV in 116 cases(71%) as the initial technique of crossing the defect. They formed the study group. This included 101 of 133 PM-VSDs(75.9%), 13 of 20 intraconal VSDs(65%) and 2 of 10(20%) post-surgical residual VSDs.(Figure 2) The proportion of cases where we elected to antegradely cannulate the VSD increased from 25.6% of the VSD device closures in the first year of the study to almost 93% of cases in the last year of the study.(Figure 3)

**Figure 2:**
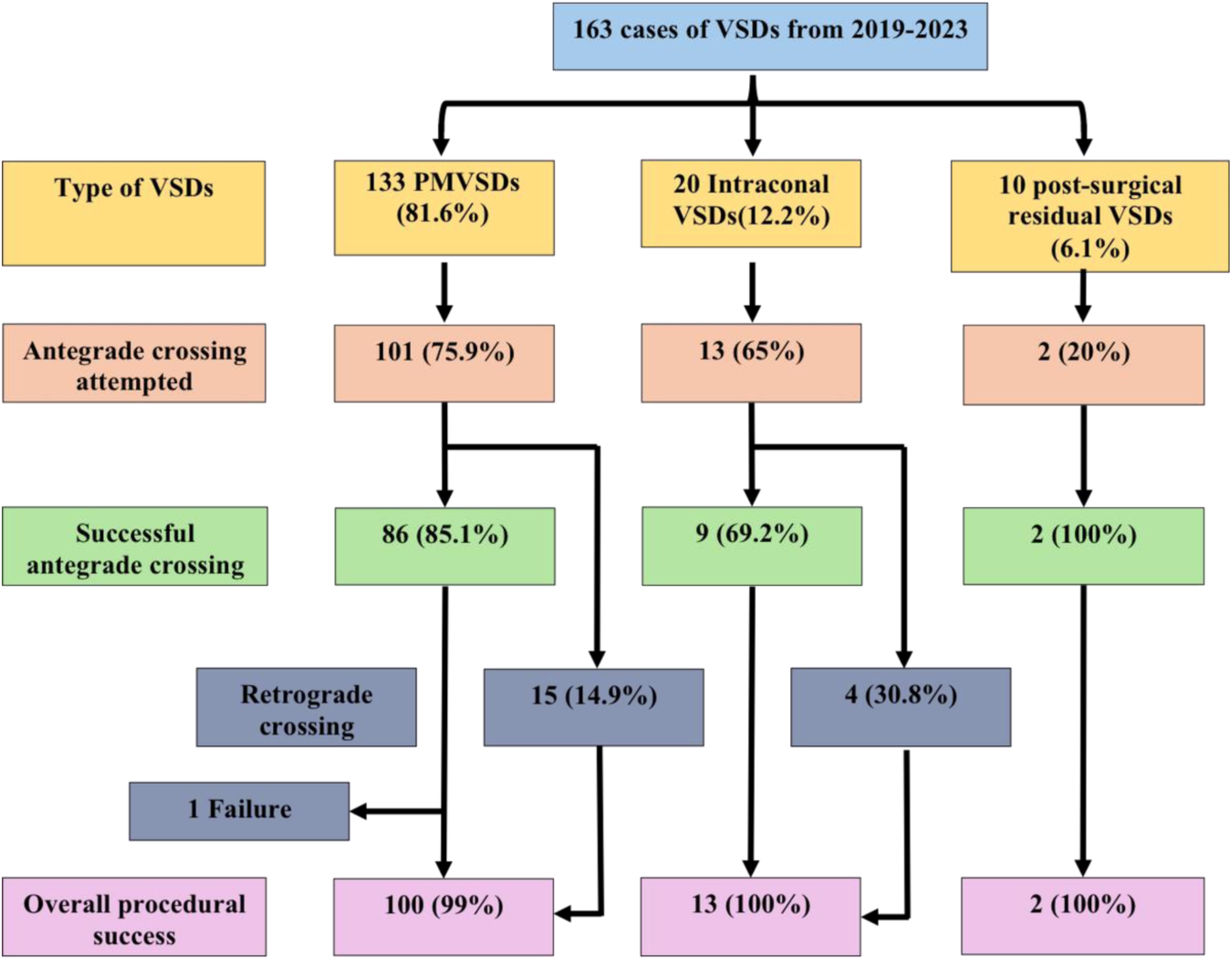
Flow chart of the study group. Among 163 device closure of ventricular septal defects near the membranous septum, an elective antegrade cannulation was adopted in 116 patients with successful transvenous procedure completion in 97 patients. The remaining 19 patients had retrograde transarterial cannulation. Overall procedural success was obtained in 115 patients, that accounted for 99.1%.

**Figure 3:**
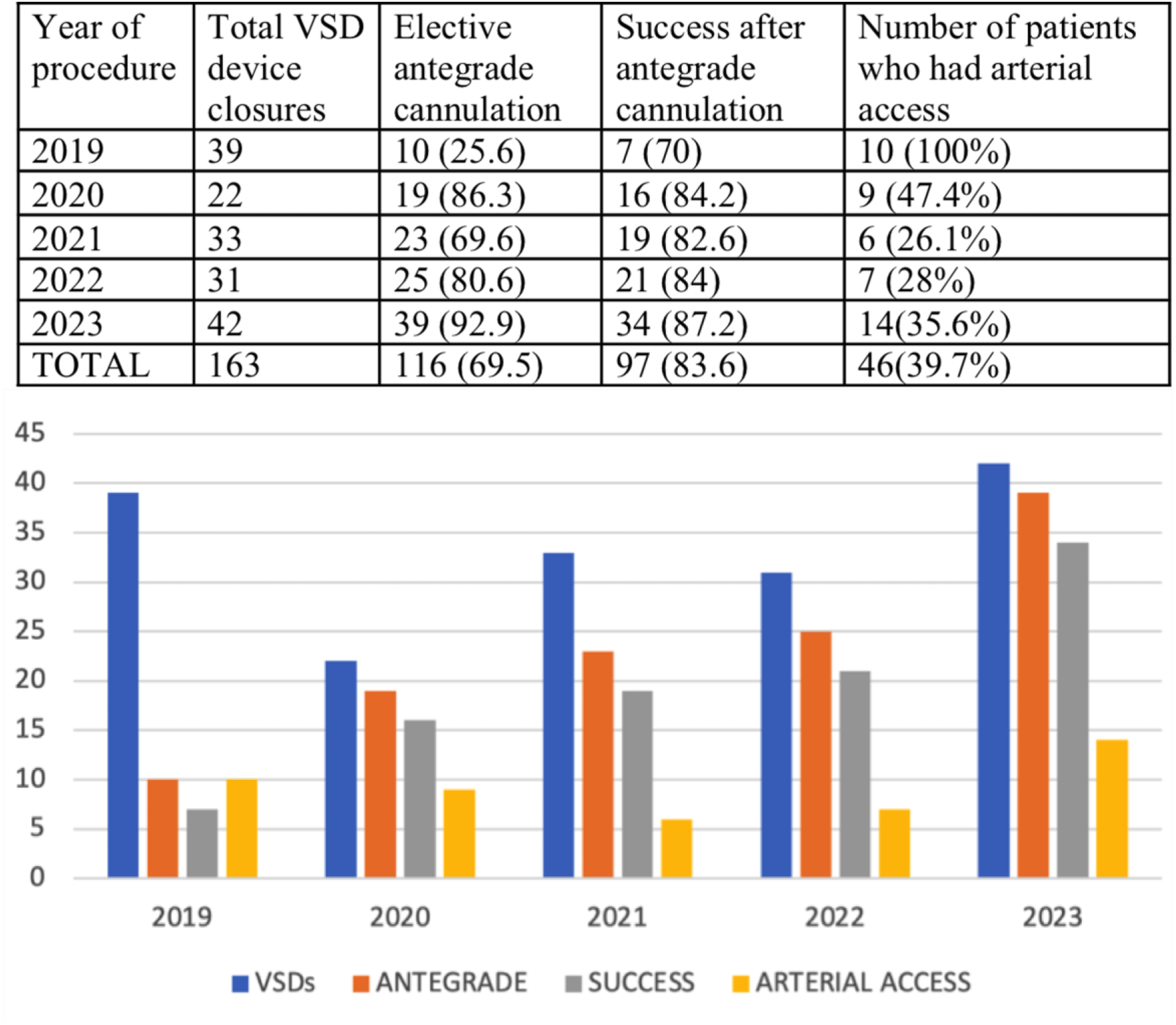
Adoption of antegrade cannulation strategy over years. While an elective antegrade cannulation was followed only in 26% of transcatheter ventricular septal defect device closures in 2019, it increased to 93% in 2023 as the confidence in the strategy improved. Even though the procedural success (83.6% overall) did not vary over the years, use of arterial access reduced over time with complete reliance on periprocedural echocardiography.

### Patient details

The median age of the cohort that included 24 infants was 55 months.(Table 1) The youngest child was one-month-old weighing 2.3kg with a 4.5mm PM-VSD and moderate hyperkinetic pulmonary hypertension and was ventilator dependent due to heart failure and refractory pneumonia. 38 patients(32.8%) had malnutrition with weight Z-score ≤ 2. Indications for closure was symptomatic state in 71 patients(61.2%) and left heart volume overload in the rest. Preprocedural electrocardiographic abnormalities included conduction disturbances in 14 patients(12.1%) and ventricular hypertrophy in 64 patients(55.2%), detailed in Table 1.

**Table 1:** Baseline patient characteristics.

|  |  |
| --- | --- |
|  | Total number of patients - 116 |
| Age (months)* | 55 (1-636) IQR 17.75-120.75 |
| Age ≤ 12 months † | 24 (20.7) |
| Age >12 years† | 21 (18.1) |
| Male gender† | 58 (50) |
| Weight (Kg) * | 15.75 (2.3 - 79) IQR 9.6-29.8 |
| Weight Z-score* | -1.25 (-7 to 1.92) IQR -2.4 to -1.25 |
| Height (cm) * | 105 (44-177) IQR 80.25-137.75 |
| Symptoms† | 71 (61.2) |
| Growth failure | 37 (31.9) |
| Dyspnoea, heart failure | 21 (18.1) |
| Frequent lung infections | 19 (16.4) |
| Pre-existing bundle branch block† |  |
| Left anterior hemiblock | 6 (5.2) |
| RBBB | 2 (1.7) |
| Bifascicular block | 6 (5.2) |
| Ventricular hypertrophy† |  |
| Biventricular hypertrophy | 32 (27.6) |
| Left ventricular hypertrophy | 32 (27.6) |
| Type of VSD† |  |
| PMVSD | 101 (87.1) |
| Type A | 9 (7.8) |
| Type B | 20 (17.2) |
| Type C | 35 (30.2) |
| Type D | 37 (31.9) |
| Intraconal VSD | 13 (11.2) |
| Post-surgical residual VSD | 2 (1.7) |
| VSD size in mm* | 6 (3-16) IQR 5-7 |
| Indexed VSD size in mm/m<sup>2</sup> BSA* | 9.2 (1.7-43.3) IQR 5.5-14.8 |
| Aortic regurgitation† |  |
| Trivial | 7 (6) |
| Mild | 2 (1.7) |
| Tricuspid regurgitation† |  |
| Mild | 21 (18.1) |
| Moderate | 4 (3.4) |
| LVID end diastole Z score* | 0.81 (-1.89 to 5.43) IQR 0.17 to 1.9 |
| LVID end systole Z score* | 0.73 (-2.45 to 4.57) IQR 0.1 to 1.8 |
| PA:Aortic systolic pressure ratio* | 0.3(0.16-1.0) IQR 0.25-0.45 |
| Pulmonary Artery Hypertension† | 80 (68.9) |
| Mild | 53 (45.7) |
| Moderate | 17 (14.7) |
| Severe | 10 (8.6) |

RBBB- Right bundle branch block, PMVSD-Perimembranous ventricular septal defect, LVID- Left ventricular internal dimension, PA-pulmonary artery, BSA-body surface area \*Data expressed in median, range and interquartile range(IQR), <sup>†</sup> Data expressed in frequency and proportion

### Echocardiographic findings

The indexed VSD size on echocardiography was 9.2(IQR 5.5-14.8) mm/m^2^. Pulmonary arterial hypertension was found in more than two-thirds of the patients, most of whom were young infants with relatively large defects. Ten patients had non-restrictive defects with near-systemic pulmonary artery pressures.(Figure 4) Type D and C defects were the most frequent and accounted for more than 60% of the 101 PM-VSDs.^[2,13]^ Mild aortic cusp prolapse in 17 patients involved RCC in 15(7 PM-VSD and 8 intraconal defects) and NCC in two PM-VSDs.(Table 2) Even though 9 included patients had baseline AR (trivial in 7 and mild in 2), two patients with mild AR and one patient with trivial AR did not show a close proximity of the VSD edges to the aortic annulus indicating an independent mechanism for AR unrelated to the VSD. Mild and moderate tricuspid regurgitation(TR) was identified in 21 and 4 patients respectively. Indirect Gerbode defect was identified as a possible contributor for TR in 14(12.1%) cases.

**Figure 4:**
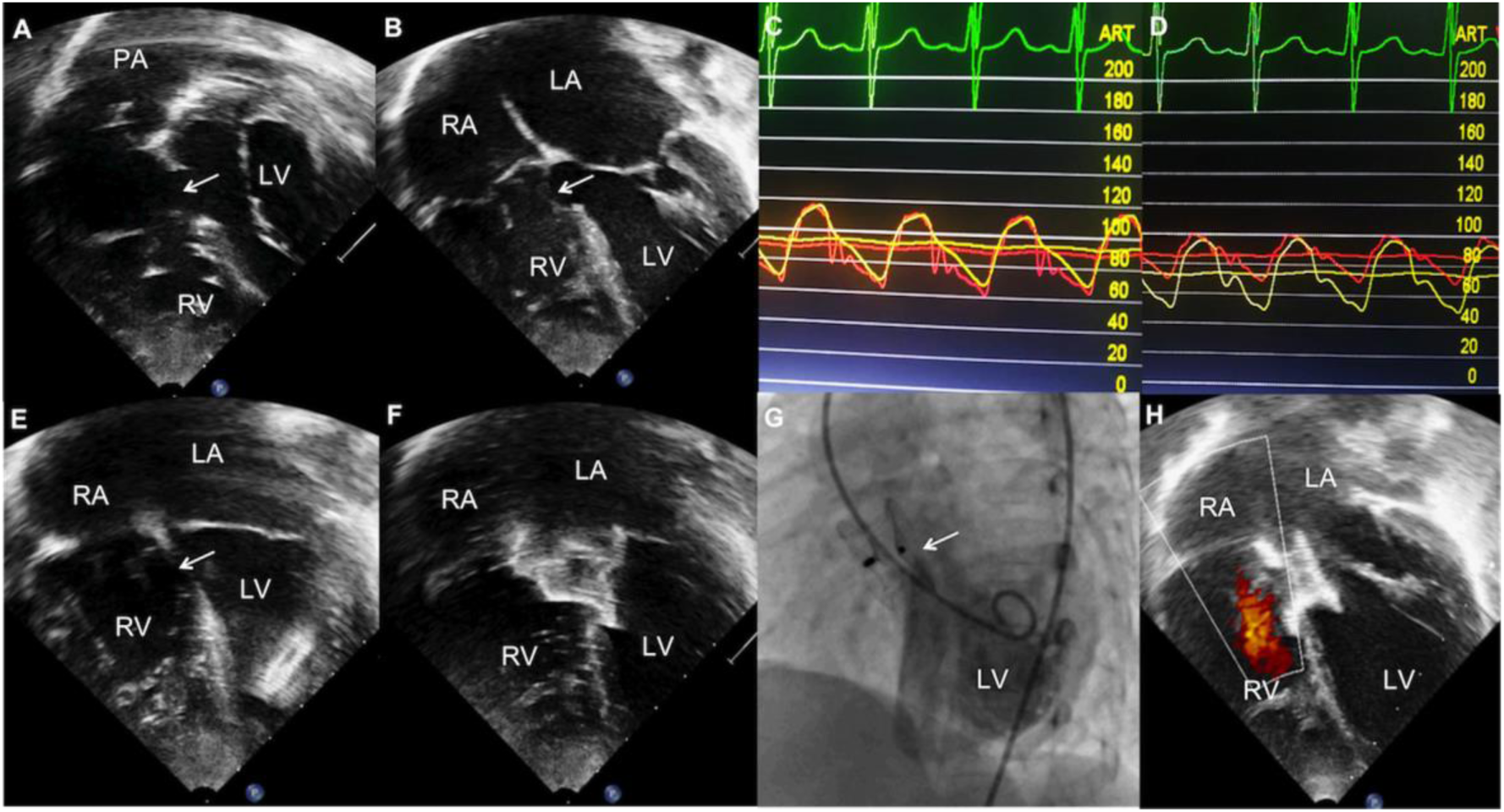
Transcatheter closure of large ventricular septal defect. Subxiphoid(A) and apical(B) views in a 2-year-old child weighing 8.5kg shows a large nonrestrictive perimembranous ventricular septal defect(arrow) between left ventricle(LV) and right ventricle(RV) with minimal left heart enlargement indicating severe pulmonary hypertension. Pulmonary artery pressure is similar to aortic pressure in room air(C) but shows acute vasodilator response to inhaled nitric oxide and oxygen(D). Antegrade cannulation of the defect(E) allows echocardiographically guided device closure(F) confirmed by fluoroscopy(G). Follow-up echocardiogram at six months(H) shows complete closure.

**Table 2:** Procedural data.

|  | N-116 |
| --- | --- |
| Anaesthesia* |  |
| Intravenous sedation | 100 (86.2) |
| Local anaesthesia | 12 (10.3) |
| Mechanical ventilation | 4 (3.4) |
| Arterial access* | 46 (39.7) |
| Successful antegrade cannulation* | 97 (83.6) |
| Device deployment (n-118)* 2<sup>nd</sup> device in 2 patients |  |
| Retrograde | 8 (6.8) |
| Antegrade | 110 (93.2) |
| First Device attempted (n-118)* |  |
| Duct occluder | 42 (35.6) |
| Amplatzer duct occluder II | 54 (45.8) |
| Konar MF occluder | 11 (9.3) |
| Muscular VSD device | 5 (4.2) |
| Piccolo occluder | 2 (1.7) |
| Others | 4 (3.4) |
| Final Device deployed (n-118) * |  |
| Duct occluder | 40 (33.9) |
| Amplatzer duct occluder II | 48 (40.7) |
| Konar MF occluder | 19 (16.1) |
| Muscular VSD device | 5 (4.2) |
| Piccolo occluder | 2 (1.7) |
| Others | 4 (3.4) |
| Need for change of device (n-118)* | 15 (12.7) |
| Insufficient oversizing | 8 (6.8) |
| Embolization | 5 (4.2) |
| Significant aortic regurgitation | 2 (1.7) |
| Procedural failure* | 1 (0.9) |
| Procedural complications* | 32 (27.6) |
| Vascular* |  |
| Transient pulse loss | 6 (5.2) |
| Deep vein thrombosis | 1 (0.9) |
| Device embolization* | 5 (4.3) |
| Rhythm complications* |  |
| Transient junctional rhythm | 4 (3.4) |
| Transient complete heart block | 2 (1.7) |
| Transient Wenckebach phenomenon | 2 (1.7) |
| Transient left bundle branch block | 1 (0.9) |
| Left anterior fascicular block | 10 (8.6) |
| Supraventricular tachycardia | 2 (1.7) |
| Residual leak post procedure (115)* | 30 (26.1) |
| Minimal | 18 (15.7) |
| Mild | 11 (9.6) |
| Moderate | 1 (0.9) |
| Duration of hospitalization (days) <sup>†</sup> | 3 IQR 2-4 |
| Procedural time (min) <sup>†</sup> | 45 IQR 30-65 |
| Fluoroscopy time (min)<sup>†</sup> | 13.3 IQR 9-21.3 |
| Dose area product (mGray.cm<sup>2</sup>)<sup>†</sup> | 8593 IQR 3910-19542 |

\*Data expressed in frequency and proportion
<sup>†</sup>Data expressed in median and interquartile range(IQR)

### Procedural data

Most procedures were performed under intravenous sedation or local anaesthesia and only 4 young infants needed general anaesthesia. Right femoral venous access was used in all cases. Among the 46 cases who required arterial access, sixteen were necessitated only after a failed antegrade cannulation. Reason for arterial access in the remaining 30 patients included hemodynamic evaluation of shunt and vascular resistance, left ventriculography for delineating multiple PM-VSD jets, coronary angiography in adults and angiographic documentation of aortic cusp prolapse. Antegrade cannulation was successful in 97(83.6%) cases including 85.1% of PM-VSDs, 69.2% of intraconal defects and 100% of post-surgical residual defects. Antegrade crossing was uniformly successful in all the 24 infants.

### Outcome after a successful antegrade cannulation

Among the 97 cases where antegrade crossing was successful, the device could be successfully deployed in first attempt from venous access in 86(88.7%) cases.(Table 3) Of the remaining 11 patients, two had residual flow through an additional fenestration, four had prolapse of the device into the RV, three had embolization of the device to pulmonary artery immediately after release and two had occurrence of AR. Residual flow through additional fenestration was successfully closed by a second device delivered retrograde in one patient with post-surgical residual defect and a second antegrade cannulation in the other with denovo type-D PM-VSD. When the undersized devices prolapsed into the RV in 4 cases, antegrade cannulation was repeated to successfully deploy another upsized device. All the three embolized devices were retrieved immediately and device was upsized after a repeat antegrade cannulation. Aortic regurgitation after ADO-II deployment related to its large LV disc in one patient was managed by retrieval, antegrade recannulation and closure with a HeartR duct occluder which protruded less on the LV side. In the other patient with AR, the initial Memopart muscular device was successfully changed to ADO-II device.

**Table 3:** Predictors of successful antegrade cannulation.

|  | Successful antegrade crossing (n-97) | Failed antegrade crossing (n-19) | P Value |
| --- | --- | --- | --- |
| Age in months<sup>†</sup> | 48 (1-600) IQR 13-99 | 119 (14-636) IQR 53-168 | 0.016<sup>‡</sup> |
| VSD size (mm)<sup>†</sup> | 6 (3-16) IQR5-7 | 6 (3-9.5) IQR5-7 | 0.3<sup>‡</sup> |
| Indexed VSD size (mm/m2)<sup>†</sup> | 9.62 (1.7-43.3) IQR 5.9-16.7 | 4.95 (3.1-11.6) IQR 3.95-10 | 0.001<sup>‡</sup> |
| Weight (Kg)<sup>†</sup> | 15 (2.3-79) IQR 8-26.5 | 25 (9.7-75) IQR 13.8-55 | 0.006<sup>‡</sup> |
| Height (cm)<sup>†</sup> | 99 (44-174) IQR 80-135 | 121 (74-177) IQR 102-159 | 0.01<sup>‡</sup> |
| Experience over years* |  |  |  |
| 2019 | 7 (70%) | 3 (30%) | 0.78<sup>§</sup> |
| 2020 | 16 (84%) | 3 (16%) |  |
| 2021 | 19 (83%) | 4 (17%) |  |
| 2022 | 21 (84%) | 4 (16%) |  |
| 2023 | 34 (87%) | 5 (13%) |  |
| Type of VSD* |  |  |  |
| PMVSD | 86 (85.1) | 15 (14.9) | 0.28<sup>§</sup> |
| Intraconal | 9 (69.2) | 4 (30.8) |  |
| Residual | 2 (100) | 0 |  |
| PMVSD type* |  |  |  |
| Type A | 7 (77.8) | 2 (22.2) | 0.26<sup>§</sup> |
| Type B | 19 (95) | 1 (5) |  |
| Type C | 27 (77.1) | 8 (22.9) |  |
| Type D | 33 (89.2) | 4 (10.8) |  |
| Aneurysm* | 60 (83.3) | 12 (16.7) | 0.91<sup>§</sup> |

PMVSD- Perimembranous ventricular septal defect
<sup>†</sup> Data expressed in median and range, \*Data expressed in frequency and proportion<sup>‡</sup> Mann Whitney U test <sup>§</sup>Chi square test

### Procedural failure

After successful antegrade cannulation and HeartR duct occluder deployment in one patient weighing 7.4kg with type D PM-VSD, moderate pulmonary hypertension and a thin inter-chordal septal aneurysm, the device embolized into the PA in the recovery room. Echocardiogram showed chordal rupture of tricuspid valve leaflet. This patient underwent surgical device retrieval, pericardial patch closure of VSD and tricuspid valve repair.

### Unsuccessful initial antegrade cannulation

Attempt to initially cannulate from venous end was unsuccessful in 19 patients, where the defect was crossed retrogradely with an arterial catheter. This was followed by a retrograde device delivery in 9 patients and AV loop formation in 10 patients. Among the 9 former patients with a retrograde device deployment, eight procedures were successful and one had AR after device placement, managed by retrieval, AV loop formation and successful antegrade delivery of the device in a deeper location avoiding a contact with the aortic valve. Among the 10 cases with AV loop formation, antegrade deployment was successful in first attempt in 7 patients. It failed in three due to device prolapse into the RV in one, embolization to PA in another and sheath kink preventing device deployment in the last patient. When the undersized device prolapsed into the RV, it was successfully upsized. Embolized device in one patient was retrieved and successfully replaced with another upsized device. Sheath kink in the last patient was finally managed by retrograde deployment of ADO-II. Overall success of TCC was achieved in 115 patients(99.1%). The initial selected device was upsized or changed in 15 patients.(Table 2)

### Factors predicting success of antegrade crossing

Various factors including patient age and size, type of VSD, indexed VSD size , experience over the years within the study period were evaluated to determine the predictors of successful antegrade crossing.(Table 3) Young age, small body size, large indexed VSD size were significantly associated with a successful antegrade cannulation from the RV side. Type of VSD, presence of septal aneurysm, year of experience did not influence the success in antegrade cannulation.

### Postprocedural complications

#### Valvular complications

Post procedural significant TR was seen in three cases, of whom surgery performed for one case associated with device embolization was described earlier. In two other patients, conservative management was adopted for moderate regurgitation as they were asymptomatic without any systemic venous congestion. While preprocedural AR was identified in 9 patients(trivial in 7 and mild in 2), 30 patients had AR on follow-up echocardiography, which was trivial in 22 and mild in eight patients. None of these patients had clinical findings of AR on examination. A similar conservative management was adopted due to asymptomatic state and lack of progress with time.

#### Arrhythmias

None of the patients needed a permanent or temporary pacemaker implantation. Transient intraprocedural complete heart block in two patients before device release recovered after reducing the tension on the delivery cable.(Table 3) Similarly left bundle branch block noted in one patient recovered within few minutes. These three patients received a short course of intravenous steroids and uneventful observation for few days. While preprocedural left anterior hemiblock, right bundle branch block and bifascicular block(combination of the above) was identified in 6,2 and 6 patients, post procedural follow-up electrocardiogram showed the above mentioned abnormalities in 9,3 and 5 patients respectively. None of the patients had persistent left bundle branch block, complete heart block or high grade conduction disturbances.

#### Device embolization

Device embolization occurred in 4 cases with PM-VSD and one with post surgical residual defect immediately after release or within hours. Three embolized ADO II devices were retrieved and replaced by upsized Konar MF occluders. Among the two others involving HeartR duct occluders, one embolization into descending thoracic aorta due to faulty deployment was managed by retrieval and a secure redeployment of the same device. The other embolization to pulmonary artery in a patient with thin interchordal septal aneurysm was sent for surgery as there was significant TR, described earlier.

#### Vascular complications

There were no immediate post-procedural venous access complications. However deep vein thrombosis was identified in one patient on fifth day when she presented with leg edema and treated with oral anticoagulation. Among the 46 cases where arterial access was used, 6 patients needed additional doses of heparin for recovery of arterial patency and none of them needed thrombolysis. There was no femoral artery occlusion on follow-up.

### Follow up data

Follow-up was complete in all 115 patients who underwent successful VSD device closure. The median follow-up duration was 25 months(1-61 months). In two infants, there was significant residual leak on follow up and elective second device closure of the residual leaks were performed after 2 and 13 months following the initial procedure. While 30 patients had residual flows on predischarge echocardiography, there was a significant reduction on the last follow-up, when 18 patients had trivial insignificant residual flows(P value 0.006). The mean LV internal dimension z-scores in diastole and systole also significantly reduced from 0.81 and 0.73 respectively before the procedure to -0.45 and 0.14 on follow-up.(Table 4)

**Table 4:** Follow-up data.

|  | N-115 | P value<sup>§</sup> |
| --- | --- | --- |
| Follow up duration (months)<sup>†</sup> | 25 (1-61) IQR 8-42 |  |
| Weight (Kgs)<sup>†</sup> | 17.9 (3.04-79) |  |
| Weight Z-score<sup>†</sup> | -0.72 (-3.3 to 1.92) IQR -1.8 to 0.4 | 0.104<sup>‡</sup> |
| LVID end diastole Z score<sup>†</sup> | -0.45 (-3.59 to 3.16) IQR -0.83 to 0.24 | <0.001<sup>‡</sup> |
| LVID end systole Z score<sup>†</sup> | 0.14 (-2.77 to 2.82) IQR -0.67 to 0.76 | 0.001<sup>‡</sup> |
| Tricuspid leak at last follow up* |  | 0.470<sup>‡</sup> |
| Mild | 28 (24.3) |  |
| Moderate | 2 (1.7) |  |
| Aortic regurgitation at follow up* |  | <0.001<sup>‡</sup> |
| Mild | 8 (6.9) |  |
| Trivial | 22 (19.1) |  |
| Bundle branch block<sup>†</sup> |  | 0.222<sup>‡</sup> |
| Left anterior hemiblock | 9 (7.8) |  |
| Bifascicular block | 5 (4.3) |  |
| RBBB | 3 (2.6) |  |
| Device closure for residual leak* | 2 (1.7) |  |
| Residual at follow up* Trivial | 18 (15.7) | 0.006<sup>‡</sup> |

LVID- Left ventricular internal diameter, RBBB-right bundle branch block
<sup>†</sup> Data expressed in median and range, \*Data expressed in frequency and proportion
<sup>‡</sup>Wilcoxon signed rank test, <sup>§</sup>P value comparing the parameters from baseline mentioned in Table 1 except for residual leak which compares immediate post procedure mentioned in Table 2 to leak during the last follow-up

## Discussion

Randomized trials and few meta-analysis establishing safety of TCC of PM-VSDs has led to an increased acceptance of the procedure in the last decade.^[6–9]^ The concept of using a transvenous access alone to antegradely cannulate the VSD from the RV and perform a successful closure seems interesting with theoretical possibility of lesser complications associated with arterial vascular sheaths and shorter procedural time. A similar approach has become the standard of care in many centres performing transcatheter PDA closure.^[15–17]^ Avoidance of AV loop formation reduces the need for additional hardware and prevents hemodynamic compromise from aorto-tricuspid splinting especially in small patients. However avoiding a good quality LV angiography that assists in fluoroscopic measurements increases the reliance on echocardiography during the antegrade cannulation strategy.

This idea of elective transvenous cannulation was tested in swine models almost 15 years ago, however was not actively pursued by interventionalists.^[18]^ Antegrade crossing was documented in only a few cases, the process of crossing was not uniform and there was no data to study the efficacy of this antegrade cannulation approach.^[19–21]^ Our present study evaluated the feasibility, efficacy and safety of antegrade transvenous crossing of PM-VSDs, intraconal VSDs and post surgical residual VSDs for the first time in literature.

### Elective antegrade cannulation strategy

Antegrade crossing of the VSD from the RV was reported in 20% to 40% of procedures in previous published studies.^[19–21]^ However, there was no data about the proportion of cases where it was attempted to determine the success rate of antegrade cannulation. An apprehension of anteroseptal tricuspid commissure acting as a curtain near the RV exit of most PM-VSDs prevented the interventionist from opting for an exclusive transvenous approach. In our centre, antegrade cannulation was attempted in 71% of transcatheter VSD device closures with increasing attempts over the period of the study from around 26% in 2019 to 93% in 2023. The procedural success of antegrade approach was 83.6% in our study indicating that this strategy was technically feasible and effective in majority of the cases.

### Factors influencing success of antegrade cannulation

Analysis of factors that predicted failure of the antegrade cannulation strategy indicated that older age, higher body surface area with relatively small defect size were associated with an unsuccessful attempt. On the contrary, younger and smaller patients with relatively larger defects had a successful antegrade cannulation. Avoidance of arterial access in small infants would reduce the access site complications.^[22,23]^ Similarly avoiding AV loop formation would simplify the procedure in infants and reduce procedural hypotension from splinting the heart valves.^[24]^ Even though the success of a procedure would be expected to improve with experience, there was no significant difference in success rates over the years in our study. This could be due to our attempts to primarily opt for antegrade cannulation in only selected PM-VSD patients in early years of study that changed to near-universal use of this strategy in almost every case of PM-VSD, intraconal VSD and post-surgical residual VSDs in the recent years.

### Benefits of antegrade cannulation

As this approach used exclusive venous access without a need for arterial sheath, complications from arterial access reported from 6.8% to as high as 33.7% from various studies were avoided.^[25–27]^ Infants prone for femoral arterial thrombosis accounted for more than 20% of our patients. Avoiding AV loop reduced the hardware, simplified the procedure and reduced the complications. This might reduce the procedural and fluoroscopic time which was 45 and 13.3 minutes respectively in our study, though this was not compared to those who had AV loop formation. Transvenous antegrade cannulation would facilitate sheath placement in LV for a device deployment in patients with aortic valve prolapse. This technique was particularly relevant in 14.6% of our patients who had aortic valve prolapse, where a retrograde transarterial delivery would preclude aortic valve monitoring and AV loop technique would necessitate manevering of the delivery system from aorta to LV before device delivery.

### Complications of antegrade crossing

As complications arising out of arterial access were inherently avoided in this antegrade transvenous cannulation technique, it was imperative to identify other potential complications related to atrioventricular nodal conduction, tricuspid and aortic valve function. Aortic valve interference would be the least expected problem with antegrade cannulation as the delivery catheter would mostly stay away from the leaflets compared to the other methods. Regarding intraprocedural evaluation of tricuspid valve function during the device deployment, antegrade approach would not allow a precise echocardiographic evaluation till the release of the device. 25 patients had preprocedural mild to moderate TR, of whom 14 patients demonstrated an indirect Gerbode phenomenon. Post-procedural follow-up echocardiogram showed TR in 30 patients, of whom two had a moderate leak. Even though this moderate TR in two patients did not result in systemic venous congestion and reach clinical significance, inability to assess tricuspid valve function before the device release from the cable remained a handicap of this technique.

### Impact on atrioventricular conduction system

The conduction tissue located in the posteroinferior margin of the VSD could be potentially injured during antegrade cannulation or device delivery. Preprocedural right bundle branch block or left anterior hemiblock identified in 12% of our patients would indicate the inherent conduction differences in these patients, especially more common in postoperative residual defects in the membranous septum. Postprocedural conduction system disturbances were identified in 18% of our patients, but majority of them were transient and clinically irrelevant. This was not significantly higher compared to the incidence ranging from 21.8% to 35.7% from the studies that meticulously identified all post-procedural arrhythmias.^[28–30]^ Left anterior hemiblock was the most common abnormality similar to the experience from other studies.^[31]^ As the surgical anatomy identified the conduction system to be closer to the LV side of the defect than the RV side, a transvenous antegrade cannulation from the RV end might not increase the possibility of post-procedural complications.^[32]^

### Limitations of the study

The number of attempts or the fluoroscopic duration required to cross the defect was not evaluated since the study was performed in an academic institution and initial attempts were done by trainees in most cases. However the median total fluoroscopic time of the procedure was 13.3 minutes only. This study also did not compare the results of antegrade approach to the conventionally followed approach that involved either a retrograde device delivery or an AV loop formation. Such a comparison would identify if antegrade technique would reduce the procedural duration or the complication rates.

## Conclusions

Antegrade cannulation of PM-VSDs, intraconal VSDs and residual VSDs in the perimembranous location followed by transvenous device delivery was found feasible in majority of the patients. Small patients with relatively large defects were the best candidates for this procedure. As this technique reduced the need for arterial sheaths and procedural complexity of AV loop formation, it was advantageous in infants. Transient conduction abnormalities identified following the procedure were clinically insignificant. An exclusive transvenous strategy might replace the currently followed retrograde device delivery or AV loop formation in future at least in small infants and young patients.

## Data Availability

The data will be available on a reasonable request to the corresponding author

## Abbreviation list

TCC: Transcatheter closure
PM-VSD: Perimembranous ventricular septal defect
VSD: ventricular septal defect
AV: arteriovenous
LV: left ventricle
RV: right ventricle
NCC: non coronary cusp
RCC: right coronary cusp
AR: aortic regurgitation
PA: pulmonary artery
ADO: Amplatzer duct occluder Konar
MF: Konar multifunction occluder

## Acknowledgements

None

## Sources of funding

None

## Disclosures

The authors have nothing to disclose, there are no conflicts of interest, there are no financial conflicts

## Notes

### Competing Interest Statement

The authors have declared no competing interest.

### Clinical Trial

This retrospective study did not require a trial ID

### Funding Statement

No funding received

### Author Declarations

Institutional review board, Madras Medical Mission, India

## References

1. Mijangos-Vázquez R, El-Sisi A, Sandoval Jones JP, et al. Transcatheter closure of Perimembranous Ventricular Septal Defects Using Different Generations of Amplatzer Devices: Multicenter Experience. J Interv Cardiol. 2020:8948249. doi: 10.1155/2020/8948249

2. Singhi AK, Sivakumar K. Echocardiographic Classification of Perimembranous Ventricular Septal Defect Guides Selection of the Occluder Design for Their Transcatheter Device Closure. J Cardiovasc Imaging. 2021;29:316–26

3. Lee SM, Song JY, Choi JY, et al. Transcatheter closure of perimembranous ventricular septal defect using Amplatzer ductal occluder. Catheter Cardiovasc Interv 2013;82:1141–6.

4. Kanaan M, Ewert P, Berger F, et al. Follow-up of patients with interventional closure of ventricular septal defects with Amplatzer duct occluder II. Pediatr Cardiol 2015;36:379–85.

5. Ho SY, McCarthy KP, Rigby ML. Morphology of perimembranous ventricular septal defects: implications for transcatheter device closure. J Interv Cardiol 2004;17:99–108.

6. Yang J, Yang L, Yu S, et al. Transcatheter versus surgical closure of perimembranous ventricular septal defects in children: a randomized controlled trial. J Am Coll Cardiol 2014;63:1159–68.

7. Butera G, Carminati M, Chessa M, et al. Transcatheter closure of perimembranous ventricular septal defects: early and long-term results. J Am Coll Cardiol 2007;50:1189–95.

8. Saurav A, Kaushik M, Mahesh Alla V, et al. Comparison of percutaneous device closure versus surgical closure of peri-membranous ventricular septal defects: a systematic review and meta-analysis. Catheter Cardiovasc Interv 2015;86:1048–56.

9. Santhanam H, Yang L, Chen Z, et al. A meta-analysis of transcatheter device closure of perimembranous ventricular septal defect. Int J Cardiol. 2018 Mar 1;254:75–83.

10. Alshahrani D, Linnane N, McCrossan B, et al. Transfemoral Perimembranous Ventricular Septal Defect Device Closure in Infants Weighing ≤ 10 kg. Pediatr Cardiol. 2023;44:1176–82.

11. Chen F, Li P, Liu S, et al. Transcatheter Closure of Intracristal Ventricular Septal Defect With Mild Aortic Cusp Prolapse Using Zero Eccentricity Ventricular Septal Defect Occluder. Circ J. 2015;79:2162–8.

12. Jiang D, Han B, Zhao L, et al. Transcatheter Device Closure of Perimembranous and Intracristal Ventricular Septal Defects in Children: Medium- and Long-Term Results. J Am Heart Assoc. 2021;10:e020417.

13. Sivakumar K. Echocardiography for interventions in congenital heart diseases: left to right shunt lesions. In: Amuthan V, Parashar S, editors. Textbook of Echocardiography. 1st ed. New Delhi: Jaypee Publishers; 2018. p.474–503.

14. Haddad RN, Daou L, Saliba Z. Device Closure of Perimembranous Ventricular Septal Defect: Choosing Between Amplatzer Occluders. Front Pediatr. 2019;7:300.

15. Baykan A, Narin N, Özyurt A, et al. Do we need a femoral artery route for transvenous PDA closure in children with ADO-I? Anatol J Cardiol. 2015;15:242–7.

16. Anil SR, Sivakumar K, Kumar RK. Coil occlusion of the small patent arterial duct without arterial access, Cardiol Young 2002;12:51–56

17. Liu J, Gao L, Tan HL, et al. Transcatheter closure through single venous approach for young children with patent ductus arteriosus: A retrospective study of 686 cases. Medicine (Baltimore). 2018;97:e11958.

18. Ratnayaka K, Raman VK, Faranesh AZ, et al. Antegrade percutaneous closure of membranous ventricular septal defect using X-ray fused with magnetic resonance imaging. JACC Cardiovasc Interv. 2009;2:224–30.

19. Kamalı H, Sivaslı Gül Ö, Çoban Ş, et al. Experiences of two centers in percutaneous ventricular septal defect closure using konar multifunctional occluder. Anatol J Cardiol 2022;26:276–285

20. Abdelmohsen GA, Gabel HA, Al-Ata JA, et al. Percutaneous closure of postoperative residual ventricular septal defects, including dehiscence of surgical patches. Cardiovasc Diagn Ther. 2023;13:710–27.

21. Kuswiyanto RB, Gunawijaya E, Djer MM, et al. Transcatheter Closure of Perimembranous Ventricular Septal Defect Using the Lifetech Konar-Multi Functional Occluder: Early to Midterm Results of the Indonesian Multicenter Study. Glob Heart 2022;17:15

22. Bansal N, Misra A, Forbes TJ, Kobayashi D. Femoral artery thrombosis after pediatric cardiac catheterization. Pediatr Cardiol. 2021;42:753–61.

23. Glatz AC, Shah SS, McCarthy AL, et al. Prevalence of and risk factors for acute occlusive arterial injury following pediatric cardiac catheterization: a large single-center cohort study. Catheter Cardiovasc Interv. 2013;82:454–62

24. Song J. Percutaneous transcatheter closure of congenital ventricular septal defects. Korean Circ J. 2023;53:134–50

25. Vitiello R, McCrindle BW, Nykanen D, et al. Complications associated with pediatric cardiac catheterization. J Am Coll Cardiol. 1998;32:1433–40.

26. Balaguru D, Dilawar M, Ruff P, Radtke WA. Early and late results of thrombolytic therapy using tissue-type plasminogen activator to restore arterial pulse after cardiac catheterization in infants and small children. Am J Cardiol. 2003;91:908–10.

27. Alexander J, Yohannan T, Abutineh I, et al. Ultrasound-guided femoral arterial access in pediatric cardiac catheterizations: A prospective evaluation of the prevalence, risk factors, and mechanism for acute loss of arterial pulse. Catheter Cardiovasc Interv. 2016;88:1098–1107.

28. Tang C, Shao S, Zhou K, et al. Complete Left Bundle-Branch Block After Transcatheter Closure of Perimembranous Ventricular Septal Defect Using Amplatzer Duct Occluder II. J Am Heart Assoc. 2022;11:e022651.

29. Jiang D, Zhang S, Zhang Y, et al. Predictors and long-term outcomes of heart block after transcatheter device closure of perimembranous ventricular septal defect. Front Cardiovasc Med. 2022;9:1041852

30. Wang C, Zhou K, Luo C, Shao S, Shi X, Li Y, Wei L, Yan S, Liu X, Hua Y. Complete Left Bundle Branch Block After Transcatheter Closure of Perimembranous Ventricular Septal Defect. JACC Cardiovasc Interv. 2019;12:1631–3.

31. Wu Z, Yang P, Xiang P, et al. Left Anterior Fascicular Block After Transcatheter Closure of Ventricular Septal Defect in Children. Front Cardiovasc Med. 2021;8:609531.

32. Milo S, Ho SY, Wilkinson JL, Anderson RH. Surgical anatomy and atrioventricular conduction tissues of hearts with isolated ventricular septal defects. J Thorac Cardiovasc Surg. 1980;79:244–55.

